# Effects of water quality, sanitation, handwashing and nutritional interventions on enteric pathogen seroconversion in rural Kenya: a cluster randomised trial

**DOI:** 10.64898/2026.10.30.26366882

**Authors:** Henry Kanyi, Richelle W. Kihoro, Huiyu Hu, Henry Emisiko, E. Brook Goodhew, Chabier Coleman, Richelle C. Charles, Edward T. Ryan, Claire Munroe, Robert Atmar, Pearl Anne Ante-Testard, Everlyn Kamau, Samantha J. Bents, Beryl S. Achando, Andrew N. Mertens, Holly N. Dentz, Gouthami Rao, Marlene K. Wolfe, Christine P. Stewart, Amy J. Pickering, Clair Null, Audrie Lin, John M. Colford, Patrick J. Lammie, Jeffrey W. Priest, Benjamin F. Arnold, Sammy M. Njenga

**Author notes:** **Corresponding author:** Benjamin F. Arnold, PhD, Professor, F.I. Proctor Foundation, University of California, San Francisco. Equal contribution (Henry Kanyi, Richelle W. Kihoro, Huiyu Hu). Equal contribution (Benjamin F. Arnold, Sammy M. Njenga).

## Abstract

**Background:** We evaluated whether water, sanitation, and handwashing (WASH) and nutrition interventions reduced enteric pathogen seroprevalence during the first two years of life among children in rural Kenya.

**Methods:** We analysed the 2×2 factorial subset of the WASH Benefits Kenya cluster-randomised trial (control, WASH, nutrition, and WASH plus nutrition). Dried blood spots were collected from all children in the birth cohort after two years of intervention and at 6 and 12 months within a longitudinal substudy. Immunoglobulin G (IgG) responses were measured using a multiplex bead assay that included 12 pathogens. The primary outcome was IgG seroprevalence at 2 years. We estimated intention-to-treat differences, comparing WASH with no WASH and nutrition with no nutrition across the factorial design. Trial registration: NCT01704105.

**Findings:** The primary analysis included 3030 children in 384 clusters (median age 23 months). Seroprevalence increased rapidly with age and exceeded 90% for norovirus, *Campylobacter*, and LT-ETEC by the final visit, with several pathogens at moderate prevalences (24% were seropositive to typhoidal *S. enterica* HlyE). Differences between intervention groups were small: lower *T. solium* (−3.3 percentage points [95% CI −6.3 to −0.2]) and higher *E. histolytica* (3.2 [0.1 to 6.3]) with WASH versus control, and lower *V. cholerae* (−1.1 [−1.8 to −0.3]) and *Shigella*/EIEC (−2.7 [−5.5 to 0.0]) with nutrition versus no nutrition. No comparison met the 5% Benjamini– Hochberg false discovery rate threshold. Secondary analyses and age-seroprevalence curves suggested modest reductions for some pathogens but increased norovirus seroprevalence around 12 months, when intervention adherence was highest.

**Interpretation:** At achieved adherence levels, household-level WASH and nutrition interventions were insufficient to reduce widespread early-life enteric pathogen exposure in this high-transmission setting, supporting the development of complementary vaccine and transmission-control strategies.

**Funding:** National Institutes of Health (R01AI166671, R01AI106878, R01AI134814). The original WASH Benefits trial was funded by the Gates Foundation (OPPGD759).

## Introduction

Enteric pathogens remain a leading cause of child morbidity and mortality, with the highest burden among young children in low-resource settings.^1^ Much of this burden is measured through clinical diarrhea and diarrhea-associated mortality, but enteric infections are also common without reported diarrhea.^2^ Repeated symptomatic and subclinical infections can contribute to acute malnutrition, linear growth faltering, impaired neurodevelopment, and other adverse outcomes.^3–8^ Enteric pathogens are transmitted through multiple pathways, including contaminated drinking water, food, hands, soil, surfaces, and contact with human and animal feces.^9–11^ For many high-burden enteric pathogens, vaccines are unavailable or not widely used in young children, with rotavirus vaccination being the main exception.^12^ Therefore, prevention largely depends on interventions that address multiple transmission pathways. Improvements in household water treatment and safe storage, sanitation improvements, and handwashing with soap (WASH) are central components of such strategies. ^13^ Nutrition interventions may also reduce susceptibility to infection and related sequelae by supporting immune function, improving micronutrient status, and promoting infant feeding practices such as exclusive breastfeeding, which can delay exposure to contaminated water and foods. ^14–17^

The WASH Benefits study included large cluster-randomised trials in rural Bangladesh and Kenya, each randomizing more than 700 clusters to individual or combined water, sanitation, handwashing, and nutrition interventions.^18–20^ In the Kenya trial, nutrition improved child linear growth, but neither the WASH interventions nor the nutrition intervention reduced caregiver-reported diarrhea, despite high diarrhea prevalence in the study population.^20^ Because efficacy trials of this scale are unlikely to be repeated often in the WASH and nutrition sectors, analyses of banked specimens can clarify which pathogens circulated and whether the interventions altered transmission despite no measured reduction in diarrhea. Stool-based analyses from WASH Benefits Kenya provided some pathogen-specific evidence, including reductions in soil-transmitted helminth infections, but there was no reduction in *Giardia* infection in response to WASH interventions.^21^ However, stool measurements capture infection only near the time of specimen collection. Infrequent stool sampling can miss infections that occur and resolve between study visits.^22^ Serologic measures could complement stool testing because pathogen-specific antibody responses integrate exposure over a longer period. In a birth cohort measured infrequently, immunoglobulin G (IgG) seroprevalence provides a more cumulative measure of prior infection and can be used to infer pathogen-specific force of infection.

In this study, we tested banked blood specimens from children enrolled in the WASH Benefits Kenya birth cohort using a multiplex IgG assay that included antigens for 12 enteric pathogens. We used these data to measure pathogen-specific seroconversion during the first two years of life and to estimate the effects of WASH and nutrition interventions on enteric pathogen transmission. We hypothesized that WASH interventions would reduce infections transmitted through environmental and person-to-person routes, and that nutrition interventions would reduce infection by lowering child susceptibility or exposure due to exclusive breastfeeding.

## Methods

### Study design and participants

WASH Benefits Kenya was a cluster-randomized, multi-arm trial that enrolled pregnant women and delivered individual and combined water, sanitation, handwashing, and nutrition interventions.18,20

The present analysis used dried blood spot (DBS) specimens collected from children in rural villages in Bungoma, Kakamega, and Vihiga counties, where most residents relied on communal water sources and unimproved sanitation, and no other WASH or nutrition interventions were ongoing. Eligibility was based on village-level environmental and infrastructural characteristics and individual-level pregnancy status within specified gestational windows. Children enrolled in the environmental enteric dysfunction (EED) substudy were visited three times during the trial for DBS specimen collection at target ages of 6, 12, and 24 months. At the final study visit (target age 24 months), DBS collection extended to all children in the birth cohort enrolled in the trial (Figure 1).

**Figure 1.**
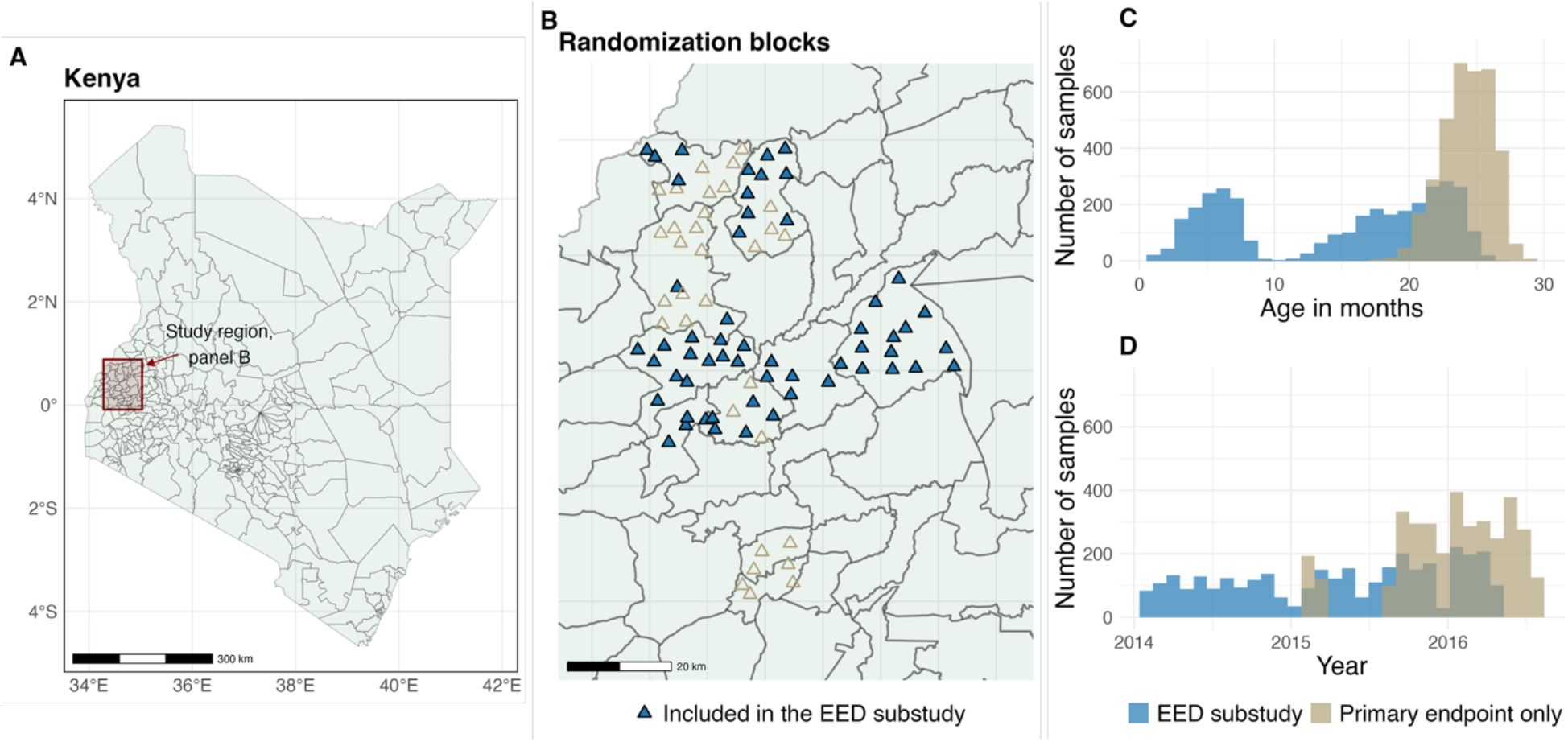
Overview of the WASH Benefits Kenya serology study. **A.** Location of the WASH Benefits Kenya study area and randomization blocks (panel B) in western Kenya. **B**. Randomization blocks contributing samples to the EED substudy (n=56) and the full study (n=88). All children in the trial were eligible for specimen collection at the final visit, so the sample set was augmented beyond the EED substudy to include remaining blocks from the full study. Each randomization block contributed 5 clusters to the analysis. **C**. Distribution of child age at blood sample collection in the EED substudy and additional children at the primary endpoint. **D**. Distribution of sample collection dates in the EED substudy and full study. EED, Environmental Enteric Dysfunction.

### Interventions

The trial included eight arms: a double-sized active control, a passive control, and six intervention arms: improved water, improved sanitation, improved handwashing, combined WASH, improved nutrition, and combined WASH plus nutrition. The water intervention promoted drinking chlorinated water (sodium hypochlorite); the sanitation intervention promoted safe disposal of faeces and provided upgraded/improved latrines with child potties and a faeces removal tool; the handwashing intervention promoted handwashing with soap and provided handwashing stations; and the nutrition intervention provided maternal/infant/young child feeding counseling plus lipid-based nutrient supplements for children from 6–24 months. The WASH arm combined the water, sanitation, and handwashing components, and WASH+N combined WASH and nutrition. In this pre-specified analysis, we analyzed IgG from children in the 2 x 2 factorial subset of trial arms: active control (hereafter control), WASH, nutrition (N), and combined WASH+N.

### Randomization and blinding

WASH Benefits Kenya was geographically matched before randomization.^18^ Within geographically matched blocks of nine clusters, clusters were randomised to trial arms; the trial included a double-sized control in each matched block, alongside single and combined intervention arms. Blinding of participants was not possible. Investigators, lab technicians, and analysts remained blinded to treatment assignments. The random allocation sequence was generated at the University of California, Berkeley using a computer-based random number generator with a reproducible seed.

### Sample collection

DBS specimens were collected from children whose caregivers provided consent for study participation and specimen collection. Trained nurses or phlebotomists collected whole-blood specimens. DBS specimens were prepared using TropBio filter paper (TropBio Pty Ltd., Queensland, Australia), with extensions pre-calibrated to hold 10 µL of whole blood. Samples were allowed to air-dry overnight at ambient temperature and were stored individually in airtight plastic bags. Samples collected at each visit were packed in larger plastic bags containing silica desiccant sachets to absorb moisture. Samples were shipped under cold chain to the laboratory in Nairobi and stored at −80°C until analysis.

### Laboratory procedures

A multiplex bead assay was used to measure IgG in DBS specimens on a Bio-Plex 200 instrument (Bio-Rad, Hercules, CA, USA) at the Kenya Medical Research Institute/Eastern and Southern Africa Center of International Parasite Control (KEMRI/ESACIPAC) laboratory. The assay was performed as described by Gwyn et al.^23^ Samples were analyzed using a single bead batch coupled at the US Centers for Disease Control and Prevention and Massachusetts General Hospital and tested at KEMRI/ESACIPAC. The multiplex panel included antigens from 12 enteric pathogens: norovirus groups I and II (GI.5, GII.3, GII.4, GII.6, GII.17); *Campylobacter* spp. (p18, p39); heat-labile toxin–producing enterotoxigenic *Escherichia coli* (LT-ETEC; LTB); *Shigella*/enteroinvasive *E. coli* (EIEC; IpaB); *Salmonella enterica* (LPS groups B and D); *S. enterica* serovars Typhi/Paratyphi (HlyE); *Vibrio cholerae* (OSP-Inaba, OSP-Ogawa); *Cryptosporidium* (Cp17, Cp23); *Giardia duodenalis* (VSP-5); *Entamoeba histolytica* (LecA); *Taenia solium* (Es33, T24); and *Strongyloides stercoralis* (NIE).

Median fluorescence intensity minus background (MFI-bg) was used as the quantitative measure of antigen-specific bound IgG in each sample. Seropositivity cutoffs for protozoan and helminth antigens were derived using receiver operating characteristic curves with known positive and negative samples (NIE, T24, Es33, Cp17, Cp23, VSP-5), or as the mean plus 3 standard deviations among 163 US residents with no known foreign travel (LecA). For bacterial and viral pathogens without known negative controls to develop seropositivity cutoffs, we used a pre-exposed cohort approach. Children in the longitudinal EED substudy who experienced a 100-fold increase in MFI-bg between measurements were identified, and their pre-boost measurements were used to characterize the seronegative distribution. The cutoff was defined as the mean plus 3 standard deviations (SD) of the pre-boost distribution. Seropositivity cutoffs were derived using samples from both the Bangladesh and Kenya trials because samples were processed in the same laboratory using the same bead couplings and reagents, and the same cutoffs were applied to both cohorts (Supplementary Figure 1).

### Outcomes

The primary outcome was pathogen-specific IgG seroprevalence measured among all children with available specimens at visit 3 (the final visit, approximate age 24 months). In this age range, children experience their first enteric infections and subsequent seroconversions.^22^ Assuming no seroreversion and negligible maternally derived IgG at visits 2 and 3, seroprevalence approximates cumulative incident seroconversion by these ages.^24^ A child was classified as seropositive if they were seropositive to any antigen measured for that pathogen. We made one exception (pre-specified): for *S. enterica*, results were reported separately for HlyE and non-typhoidal serotypes (LPS groups B and D) due to the potential for HlyE to inform typhoid vaccine delivery.^25^

Prespecified secondary outcomes included seroprevalence at visit 2 (approximately 12 months) among children in the EED substudy and the count of seropositive enteric pathogens per sample (0-12). Because maternally derived IgG could contribute to seropositivity at visit 1, these measurements were used to describe age-seroprevalence curves and inform seropositivity cutoffs, but not for formal comparisons between intervention groups.

### Statistical analyses

Sample size was determined by the availability of banked specimens. Minimum detectable effects were estimated assuming 80% power, a two-sided significance level of 0.05, an intracluster correlation coefficient of 0.15, and 90 clusters per comparison group with an average of eight children per cluster for the pooled factorial comparisons. At an assumed seroprevalence of 50% in the control group, the detectable relative reduction was approximately 20%, equivalent to a 10-percentage-point absolute reduction. These calculations were based on the smaller EED substudy and did not account for potential gains in precision from geographic matching.^26^

The primary analysis was intention-to-treat and focused on the 2 x 2 factorial design, estimating superiority comparisons on the two margins: improved WASH versus no WASH and improved nutrition versus no nutrition. Analyses followed a prespecified statistical analysis plan finalized before serology results were examined. For seroprevalence at visit 3, we estimated risk differences using a linear-binomial regression model with cluster-robust standard errors to allow for correlation within clusters. Models included indicators for geographically matched block and separate indicators for WASH and nutrition. Additive interaction was assessed using a WASH × nutrition interaction term; if there was no evidence of interaction (*P* > 0.1), arms were pooled across the factorial design to estimate marginal effects of WASH and nutrition. Two-sided permutation *P*-values were estimated by permuting treatment assignments within matched blocks (10,006 replicates) and using the regression coefficients for WASH and nutrition as test statistics. Randomization test *P*-values were then adjusted for multiple comparisons across the 12 pathogens using the Benjamini–Hochberg procedure (5% false discovery rate).

The analysis was repeated among children measured at visit 2 in the EED substudy, but since the substudy did not retain geographic matching when selecting clusters, the analysis did not include indicators for matched blocks. For the count of seropositive pathogens (range 0-12), we summarized the distribution by group at visit 3 and estimated differences using linear regression with indicators for WASH and nutrition, matched-block indicators, and cluster-robust standard errors. Primary analyses were complete-case analyses.

Visit-specific estimates included one measurement per child with an available pathogen-specific result at that visit. Age-seroprevalence curves were estimated using locally weighted regression (LOESS) of binary seropositivity on observed age at specimen collection for each pathogen and by WASH or nutrition assignment, using available child-visit measurements across all three visits.

Analyses were conducted in R (version 4.5) and full replication files are available (10.5281/zenodo.23673944).

### Ethics and registration

The WASH Benefits Kenya trial was registered with ClinicalTrials.gov (NCT01704105), and the protocol was approved by ethics committees at the University of California, Berkeley, Stanford University, and the Kenya Medical Research Institute. IgG analyses of banked specimens were approved by the institutional review boards at University of California, San Francisco (#22-36722) and KEMRI (SERU #4646) under consent for future use. Written informed consent was provided by the parents of participating children. The consent process was undertaken in local languages and included provisions for non-literate participants. No additional harms were reported for this substudy.

## Results

### Study population and intervention adherence

Of the 702 clusters randomised in the WASH Benefits Kenya trial, 391 were in the four factorial arms included in this serology substudy: 158 control, 76 WASH, 78 nutrition, and 79 WASH + nutrition. In total, 5,046 DBS samples from 3,305 children were assayed across the three visits (Supplementary Figure 2). Of the 3,305 children assayed at any visit, 3,030 contributed to the primary analysis: 1,247 control (156 clusters), 582 WASH (75 clusters), 582 nutrition (77 clusters), and 619 WASH + nutrition (76 clusters). Baseline household characteristics were similar across arms and factorial comparisons (marginal and conditional) (Table 1 and Supplementary Table 3): fewer than 20% of households owned an improved latrine and fewer than 15% had soap available near a handwashing location. In the WASH groups, detectable chlorine in stored water increased to 42% at year 1 and then declined to 19% at year 2, and handwashing locations with water and soap increased to 77% at year 1 and then declined to 20% at year two; improved latrine access remained high: 89% at year 1 and 81% at year 2. Reported supplement consumption in the nutrition groups was 95% of expected consumption at year 1 and 115% at year 2, indicating reported consumption above the expected quantity at year 2 (Supplementary Table 7).

**Table 1.** Baseline characteristics by WASH and nutrition margins of the 2×2 factorial design in the WASH Benefits Kenya trial. Data are mean (SD) or n (%). Percentages are calculated among participants with nonmissing data for each characteristic. Supplementary Table 2 includes arm-specific summaries.

| Characteristic | No WASH<br>(N=1829) | WASH<br>(N=1201) | No Nutrition<br>(N=1829) | Nutrition<br>(N=1201) |
|---|---|---|---|---|
| **Maternal** |  |  |  |  |
| Age (years) | 26.5 (6.3) | 26.5 (6.3) | 26.5 (6.3) | 26.5 (6.3) |
| Completed at least primary education | 864 (47%) | 550 (46%) | 850 (47%) | 564 (47%) |
| Height (cm) | 160 (5.8) | 160 (5.9) | 160 (5.9) | 160 (5.9) |
| Study child is firstborn | 356 (19%) | 216 (18%) | 326 (18%) | 246 (21%) |
| **Paternal** |  |  |  |  |
| Completed at least primary education | 1057 (62%) | 686 (61%) | 1045 (61%) | 698 (62%) |
| Works in agriculture | 776 (44%) | 527 (46%) | 783 (45%) | 520 (45%) |
| **Household** |  |  |  |  |
| Number of households per compound | 1.6 (1.1) | 1.7 (1.1) | 1.7 (1.1) | 1.7 (1.2) |
| Number of people per compound | 8.3 (5.7) | 8.4 (5.3) | 8.2 (5.3) | 8.5 (5.8) |
| Number of children <18 years in household | 2.9 (2) | 3 (2) | 2.9 (2) | 3 (2) |
| Has electricity | 123 (7%) | 89 (7%) | 116 (6%) | 96 (8%) |
| Has a cement floor | 91 (5%) | 61 (5%) | 91 (5%) | 61 (5%) |
| Owns a mobile phone | 1480 (81%) | 969 (81%) | 1469 (80%) | 980 (82%) |
| Owns a motorcycle | 180 (10%) | 89 (7%) | 165 (9%) | 104 (9%) |
| **Drinking water** |  |  |  |  |
| Primary drinking water source is improved* | 1359 (75%) | 880 (74%) | 1342 (74%) | 897 (75%) |
| One-way walking time to primary water source (min) | 10.7 (11.6) | 10.8 (12.8) | 10.6 (12.1) | 11 (11.9) |
| Reported treating stored water | 184 (12%) | 135 (14%) | 193 (13%) | 126 (13%) |
| **Sanitation** |  |  |  |  |
| Own any latrine | 1528 (84%) | 1016 (85%) | 1529 (84%) | 1015 (85%) |
| Access to improved latrine | 281 (16%) | 175 (15%) | 292 (17%) | 164 (14%) |
| Human faeces observed in the compound | 161 (9%) | 109 (9%) | 152 (8%) | 118 (10%) |
| **Handwashing location** |  |  |  |  |
| Has water within 2 m of handwashing location | 457 (25%) | 323 (27%) | 468 (26%) | 312 (26%) |
| Has soap within 2 m of handwashing location | 154 (8%) | 149 (12%) | 177 (10%) | 126 (11%) |
| **Food security** |  |  |  |  |
| Prevalence of moderate-to-severe household hunger | 187 (10%) | 131 (11%) | 186 (10%) | 132 (11%) |

**Figure 2.**
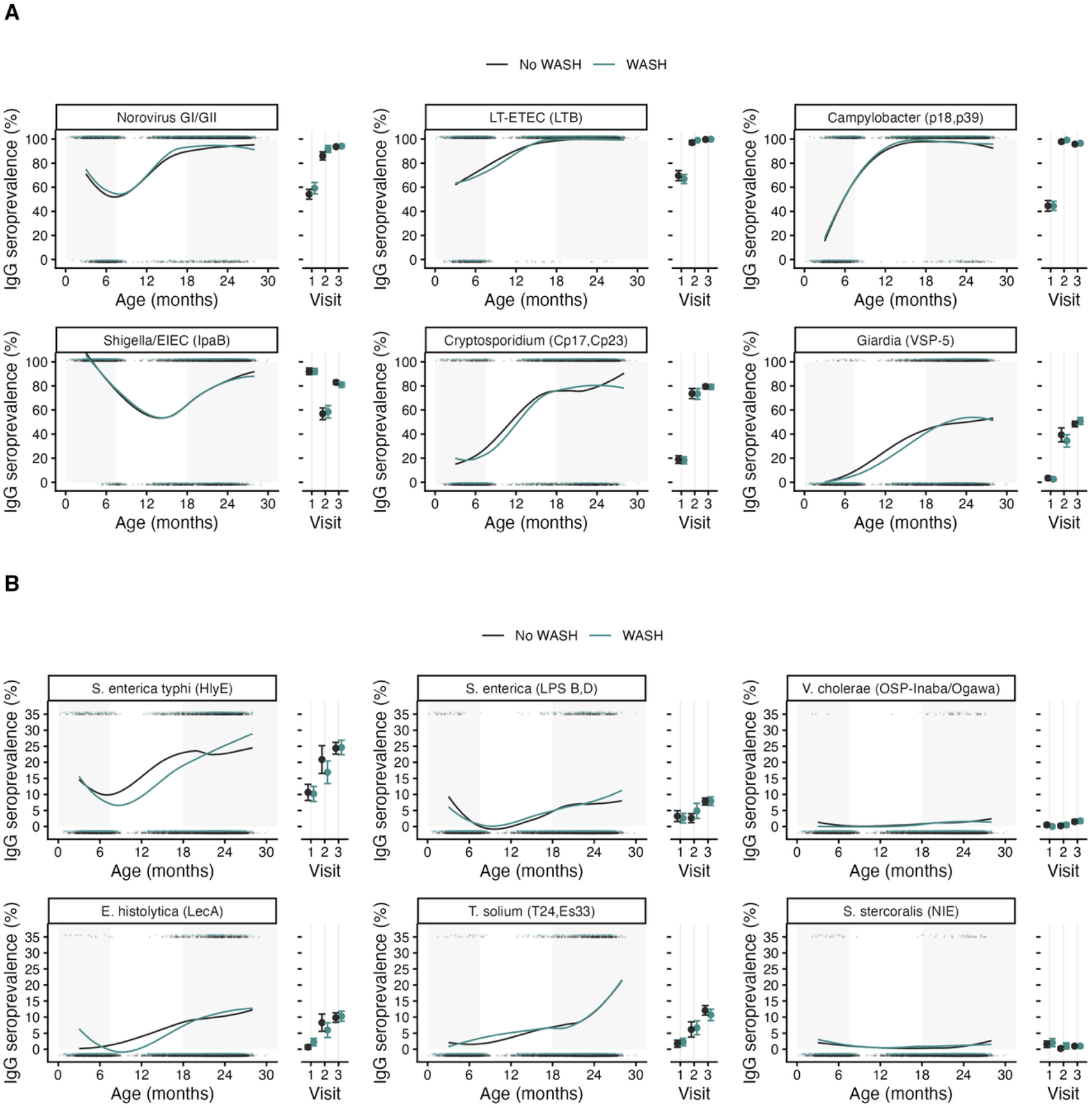
Age-dependent seroprevalence for enteric pathogens by WASH treatment in the WASH Benefits Kenya trial. **A.** Higher transmission pathogens. **B**. Lower transmission pathogens. The only difference between panels is the Y-axis range, to better show the range of the data. Lines age-seroprevalence estimated by treatment group using locally weighted regression over individual data (points), trimmed to ages 4-28 months to avoid edge effects. Points and error bars to the right of each panel show treatment-specific marginal average seroprevalence and 95% confidence intervals by study visit (1, 2, 3). Gray shading shows child age ranges of visits 1 and 3. Estimates summarized by the nutrition treatment margin are in Supplementary Figure 5.

### Enteric pathogen seroprevalence by age

Enteric pathogen seroprevalence increased rapidly during the first two years of life (Figure 2). Seroprevalence for *Campylobacter*, LT-ETEC, and norovirus reached 90-95% by age 18 months. At the final visit (median age 23 months), seroprevalence remained very high for LT-ETEC and *Campylobacter* (>95%) and high for norovirus (94%), *Shigella/*EIEC (>80%), and *Cryptosporidium* (80%) across study groups. These patterns indicate a substantial burden of exposure to enteric pathogens in early childhood in this setting. The age-seroprevalence curves suggested slightly later exposure among children who received WASH for *Giardia, S. enterica* serovars Typhi/Paratyphi, and *E. histolytica*, but intervention-specific seroprevalence generally converged by the final visit.

### Primary outcome: seroprevalence at final visit

At visit 3 (median age 23 months), IgG seroprevalence varied across the 12 enteric pathogen outcomes. For pathogens without evidence of WASH × nutrition interaction, marginal risk differences on the WASH margin were small and compatible with no effect (Figure 3A; Supplementary Table 4). On the nutrition margin, *V. cholerae* seroprevalence was lower with nutrition than without nutrition (1.1% vs 2.2%; difference, -1.1 percentage points, 95% CI -1.8 to -0.3; permutation *P* = 0.02), and *Shigella*/EIEC seroprevalence was modestly lower (80.6% vs 83.3%; difference, -2.7 percentage points, 95% CI -5.5 to 0.0; permutation *P* = 0.08; Figure 3B). We found moderate evidence of additive WASH × nutrition interaction (*P* < 0.1) for *E. histolytica* and *T. solium*. Compared with control, single-arm WASH was associated with higher *E. histolytica* seroprevalence (+3.2 percentage points, 95% CI 0.1 to 6.3) and lower *T. solium* seroprevalence (-3.3 percentage points, 95% CI -6.3 to -0.2). Nutrition showed a similar-magnitude difference for *E. histolytica* (+3.2 percentage points, 95% CI 0.0 to 6.5), but a smaller difference for *T. solium* that was compatible with no effect (-1.3 percentage points, 95% CI -4.3 to 1.7; Figure 3). After Benjamini–Hochberg adjustment across the 12 pathogen outcomes, no WASH or nutrition comparison met the 5% false discovery rate threshold.

**Figure 3.**
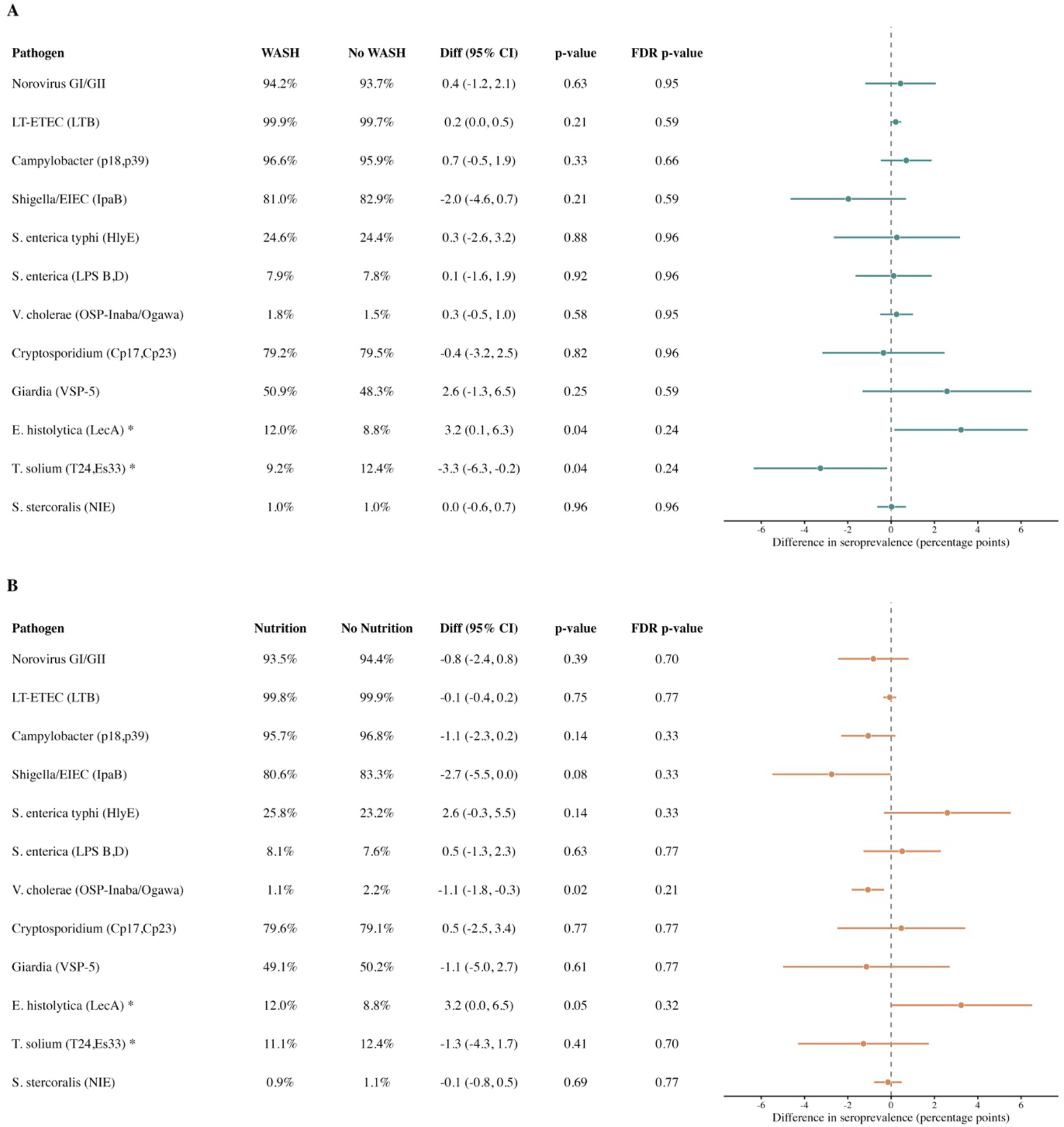
Pathogen-specific seroprevalence and differences between treatment groups at the 24-month visit in the WASH Benefits Kenya trial. Median (IQR) age of children was 23 (22, 25) months. **A.** Seroprevalence in clusters that received improved WASH versus those that did not receive improved WASH. **B**. Seroprevalence in clusters that received improved nutrition versus those that did not receive improved nutrition. Each panel includes differences between groups. In the case of moderate evidence of interaction between WASH and nutrition for *E. histolytica* and *T. solium* (*P* < 0.1 for interaction), the figure includes conditional effects where the comparison is between the single intervention WASH or Nutrition and the control group (*). Treatment-specific means and sample sizes by arm are presented in Supplementary Table 4. CI, confidence interval; FDR, false discovery rate.

### Secondary outcomes

Among EED substudy children measured at visit 2 during the period of highest WASH adherence (Supplementary Table 7), we found modest evidence of reductions in *Giardia* with WASH alone and *E. histolytica* with WASH alone and nutrition alone versus control, but higher norovirus GI/GII seroprevalence in the intervention arms than in control (Figure 4). Additionally, there was some evidence for lower LT-ETEC seroprevalence with nutrition than without nutrition (Figure 4B). Between-group differences at 12 months were broadly consistent with small shifts in the age-seroprevalence curves, suggesting a delay in seroconversion in the case of marginal reductions due to intervention, or slightly earlier seroconversion in the case of norovirus (Figure 2, Supplementary Figure 5).

**Figure 4.**
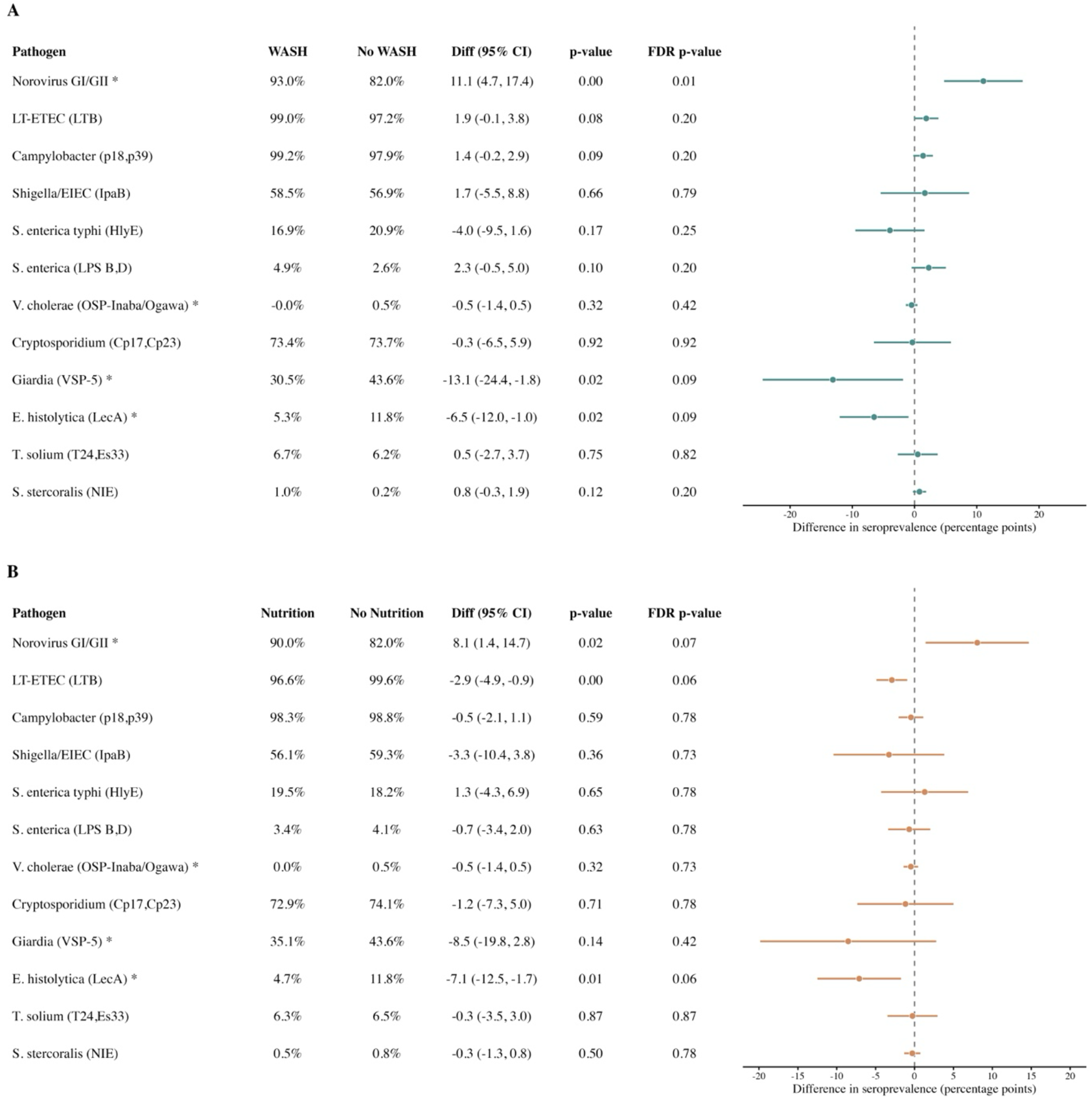
Pathogen-specific seroprevalence and differences between treatment groups at the 12-month visit in the WASH Benefits Kenya trial. Median (IQR) age of children was 17 (15, 18) months. **A.** Seroprevalence in clusters that received improved WASH versus those that did not receive improved WASH. **B**. Seroprevalence in clusters that received improved nutrition versus those that did not receive improved nutrition. Each panel includes differences between groups. In the case of moderate evidence of interaction between WASH and Nutrition for norovirus, *V. cholerae, Giardia*, and *E. histolytica* (*P* < 0.1 for interaction), the figure includes conditional effects where the comparison is between the single intervention WASH or nutrition and the control group (*). CI, confidence interval; FDR, false discovery rate.

At the final visit, most children were seropositive to at least five of the 12 pathogens. The distributions overlapped substantially between WASH and no-WASH groups and between nutrition and no-nutrition groups, with no evidence for reductions due to intervention (Supplementary Figure 6).

## Discussion

This study shows that children in rural western Kenya experienced intense enteric pathogen exposure during the first two years of life. IgG seroprevalence increased rapidly and most children were seropositive for *Campylobacter*, LT-ETEC, and norovirus by 24 months. Seroprevalence at the final visit was also high for *Shigella*/EIEC and *Cryptosporidium*. Neither the WASH nor nutrition intervention clearly reduced final-visit IgG seroprevalence after correction for multiple comparisons. Secondary analyses suggested differences for selected pathogens at visit 2. The lack of clear effect on seroprevalence at visit 3 is consistent with the trial’s primary outcomes, in which neither intervention reduced caregiver-reported diarrhoea, and nutrition-containing arms modestly improved linear growth.^20^ Environmental sampling in WASH Benefits Kenya found that combined WASH interventions improved stored drinking water quality but did not reduce *E. coli* contamination on children’s hands or toys.^27^ Similarly, in the SHINE trial in Zimbabwe, WASH and infant and young child feeding interventions did not significantly reduce individual enteropathogen prevalence after adjustment for multiple comparisons, although WASH modestly reduced the number of parasites detected.^28^ The MapSan trial in urban Mozambique found limited effects of improved compound WASH interventions on enteropathgen infection over 24 months.^29^ The limited effects may reflect continued exposure through pathways inadequately addressed by the household-level interventions, including animal faeces, which were not specifically targeted, contaminated food, and contamination outside the household. These findings support calls for more intensive WASH approaches that address multiple transmission pathways at household and community levels.

Several factors may explain the limited intervention effects at the final visit. Transmission may have been high enough that household- or compound-level WASH improvements were insufficient to substantially reduce child exposure. Children could still encounter pathogens through contaminated food, soil, surfaces, animal faeces, or contact outside the treated compound. ^9,11^ In addition, chlorination of drinking water would not be expected to protect equally against all pathogens, especially protozoa such as *Cryptosporidium* and *Giardia*.^21^ Adherence to some WASH components, especially water treatment and handwashing, declined after the first year, potentially attenuating effects at the final visit (Supplementary Table 7). ^20^

Although final-visit estimates showed no clear intervention effects, age-seroprevalence curves suggested small shifts compatible with slightly later increase in seroprevalence for selected pathogens. These shifts were most notable for *Shigella*/EIEC, LT-ETEC, *V. cholerae*, and *E. histolytica* among children assigned to nutrition, as well as *Giardia, E. histolytica and* typhoidal *Salmonella* among children assigned to WASH. However, the curves largely converged by age two years, suggesting that any effects were more consistent with transient delays in exposure than with sustained reductions in cumulative exposure. Delaying early-life infections could be clinically relevant,^30^ though we acknowledge a modest shift in the age-seroprevalence curve does not establish a clinical benefit, and neither intervention reduced caregiver-reported diarrhea in the trial.

Consistent with this interpretation, differences between intervention groups were most evident at visit 2, when WASH adherence was higher. Among children in the EED substudy, *Giardia* seroprevalence was lower in the WASH arm than in control (−13.1 percentage points, 95% CI −24.4 to −1.8), and *E. histolytica* seroprevalence was lower with WASH and nutrition than in control after one year of intervention. On the nutrition margin, LT-ETEC seroprevalence was lower with nutrition than without nutrition (−2.9 percentage points, 95% CI −4.9 to −0.9; false discovery rate–adjusted P = 0.06). These reductions were not evident at the final visit. *E. histolytica* estimates reversed direction at the final visit, with higher seroprevalence for WASH and nutrition than in control; this pattern does not support a simple delay in exposure. At the final visit, WASH was associated with 3.3 percentage points lower T. solium seroprevalence than control, whereas nutrition showed no clear effect; the WASH comparison did not meet the 5% false discovery rate threshold. Age-seroprevalence curves showed slightly later increases among children who received WASH for *Giardia* and *S. enterica* serovars Typhi/Paratyphi, but convergence between intervention groups by the final visit. For these pathogens, the curves were compatible with transient delays in exposure rather than sustained reductions in cumulative exposure. Norovirus was an exception: seroprevalence at visit 2 was higher in all three intervention arms than in control. The common direction of the norovirus and *E. histolytica* comparisons at visit 3 could partly reflect chance variation in their shared control group though this unexpected finding may warrant further investigation. The lower *V. cholerae* seroprevalence with nutrition at visit 3 could reflect differences in exposure or susceptibility to infection but since it was not significant after correction for multiple testing we cannot rule out chance.

Nearly one-quarter of children were HlyE seropositive at the end of follow-up, indicating substantial exposure to typhoidal *Salmonella* before typhoid conjugate vaccine (TCV) introduction.

These data provide a historical HlyE-based pre-TCV benchmark for evaluating population-level changes following Kenya’s national TCV introduction in 2025. Recent serology studies in Bangladesh, Malawi, and Nepal found that HlyE-based seroincidence correlated with observed enteric fever incidence across ages and sites, supporting its use in population-level surveillance.^31^ Other studies have documented typhoidal *Salmonella* transmission in Kenya, including HlyE-based seroincidence estimates of 3.6 and 37 per 100 person-years in western and coastal Kenya, respectively.^32^ These rates are not directly comparable with the seroprevalence measured here, but assuming no seroreversion 24% cumulative incidence between ages 6-24 months translates to an incidence rate of 16 per 100 person-years. Although enteric fever is often studied in urban settings, our findings suggest substantial exposure also occurs in rural populations. However, HlyE seropositivity reflects prior immune responses, not clinically confirmed enteric fever.

This study has multiple strengths. IgG testing was embedded in a large cluster-randomised trial, reducing confounding and supporting an intention-to-treat interpretation. A further strength is the use of objective, sensitive, pathogen-specific measures of infection. Serology can detect infections that occur between study visits: antibody levels integrate exposure over time and can provide a more complete picture of cumulative infection than periodic stool testing alone. However, several limitations remain. First, WASH adherence was imperfect and declined after the first year of the trial. Adherence indicators for sanitation, handwashing, and nutrition were greater than 70% at year 1, but by year 2 handwashing and water treatment adherence fell to less than 25%, which could have attenuated the treatment effects, especially for the pathogens transmitted through water or hand to mouth. ^20^ Second, IgG responses reflect prior pathogen-specific infections but, without further characterization, may not reflect the number, timing, or clinical severity of infections. Characterizing IgG kinetics with time since infection may help estimate timing for less common pathogens, such as typhoidal *S. enterica* or *V. cholerae*,^25,33^ but it remains unclear if similar modeling approaches would work for higher transmission pathogens with repeat infections and frequent IgG boosting typical of many pathogens studied here. Variation in antibody kinetics, cross-reactivity, and seroreversion may also affect interpretation of IgG. The nutrition intervention and breastfeeding might also have altered the IgG response to infection, so differences in seroprevalence may not solely reflect differences in pathogen exposure. Finally, seroprevalence approached saturation for LT-ETEC, *Campylobacter*, and norovirus, creating ceiling effects that limited detection of modest differences, and correction for multiple comparisons further reduced sensitivity to small pathogen-specific effects.

In conclusion, children in rural western Kenya experienced widespread exposure to diverse enteric pathogens by age 2 years. At the adherence levels achieved, neither WASH nor nutrition clearly reduced seroprevalence after correction for multiple comparisons, although age patterns were compatible with modest, transient shifts in seroprevalence for selected pathogens. Large-scale multiplex IgG testing represents an efficient platform to characterize cumulative exposure to many enteric pathogens from a single blood spot, identify transmission patterns difficult to capture through periodic stool testing, and establish benchmarks for future enteric vaccine and transmission-control evaluations.

## Data Availability

De-identified data underlying the reported results, the data dictionary, and R analysis code will be publicly available in the Zenodo repository (DOI: 10.5281/zenodo.23673944) upon publication.

## Contributors

Conceptualization: HK, RWK, HH, PJL, JWP, BFA, SMN. Data curation: RWK, HH, PAT, EK, SJB, ANM, BFA. Formal analysis: HH, PAT, EK, SJB, BFA. Funding acquisition: ETR, CPS, AJP, CN, JMC, PJL, JWP, BFA, SMN. Investigation: HK, RWK, HH, HE, PAT, BSA, HND,

GR, MKW, BFA, SMN. Methodology: HH, PAT, EK, SJB, JWP, BFA, SMN. Project administration: HK, RWK, BSA, HND, GR, MKW, AL. Resources: HK, RWK, HE, EBG, CC, RCC, ETR, CM, RLA, JWP. Software: HH, PAT, EK, SJB, BFA. Supervision: HK, EBG, CC, CPS, AJP, CN, AL, JMC, BFA, SMN. Validation: RWK, HE, EBG, CC, RCC, CM. Visualization: HH, PAT, SJB. Writing – original draft: HK, RWK, HH, BFA, SMN. Writing – review & editing: All authors.

## Data sharing

De-identified data underlying the reported results, the data dictionary, and R analysis code will be publicly available in the Zenodo repository (DOI: 10.5281/zenodo.23673944) upon publication. The study protocol and statistical analysis plan will also be available in the repository.

## Acknowledgments

The authors are grateful to Dr. Diana Martin, Gretchen Cooley, and Dr. Jan Vinje at the US Centers for Disease Control and Prevention for their contributions to the multiplex IgG testing. We thank William Petri (University of Virginia) and Joel Herbein (TechLab) for providing the LecA protein.

## Declaration of generative AI and AI-assisted technologies

Claude Opus 5.5 (Anthropic) assisted with Supplementary Table 7, R code review, and data/code documentation. The authors reviewed all outputs and take full responsibility.

