## Supplementary Material for "Effects of water quality, sanitation, handwashing and nutritional interventions on enteric pathogen seroconversion in rural Kenya: a cluster randomised trial"

**Supplementary Figure 1: Luminex IgG response (MFI-bg) to enteric pathogens during visits 1 and 2 in the WASH Benefits Kenya environmental enteric dysfunction substudy.** Children were median ages 3 months (visit 1) and 14 months (visit 2) at the time of measurement. Vertical lines mark seropositivity cutoffs derived using external samples (solid) for protozoan and helminth pathogens, and from longitudinal boosting in a pre-exposed cohort approach (dashed).

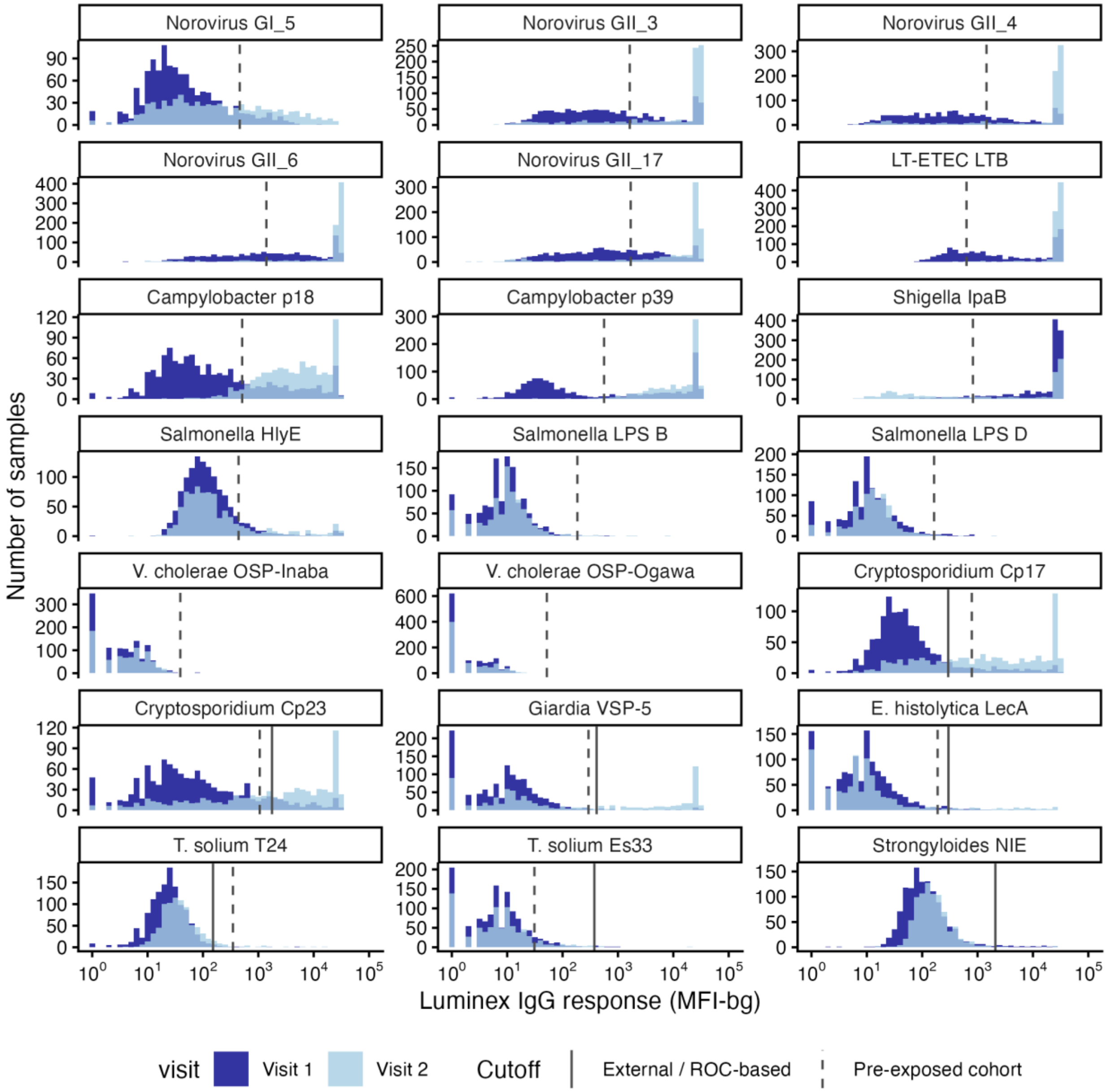

**Supplementary Figure 2. CONSORT diagram for the WASH Benefits Kenya trial serology study**. DBS, dried blood spot; EED, environmental enteric dysfunction; WASH, water, sanitation, and handwashing.

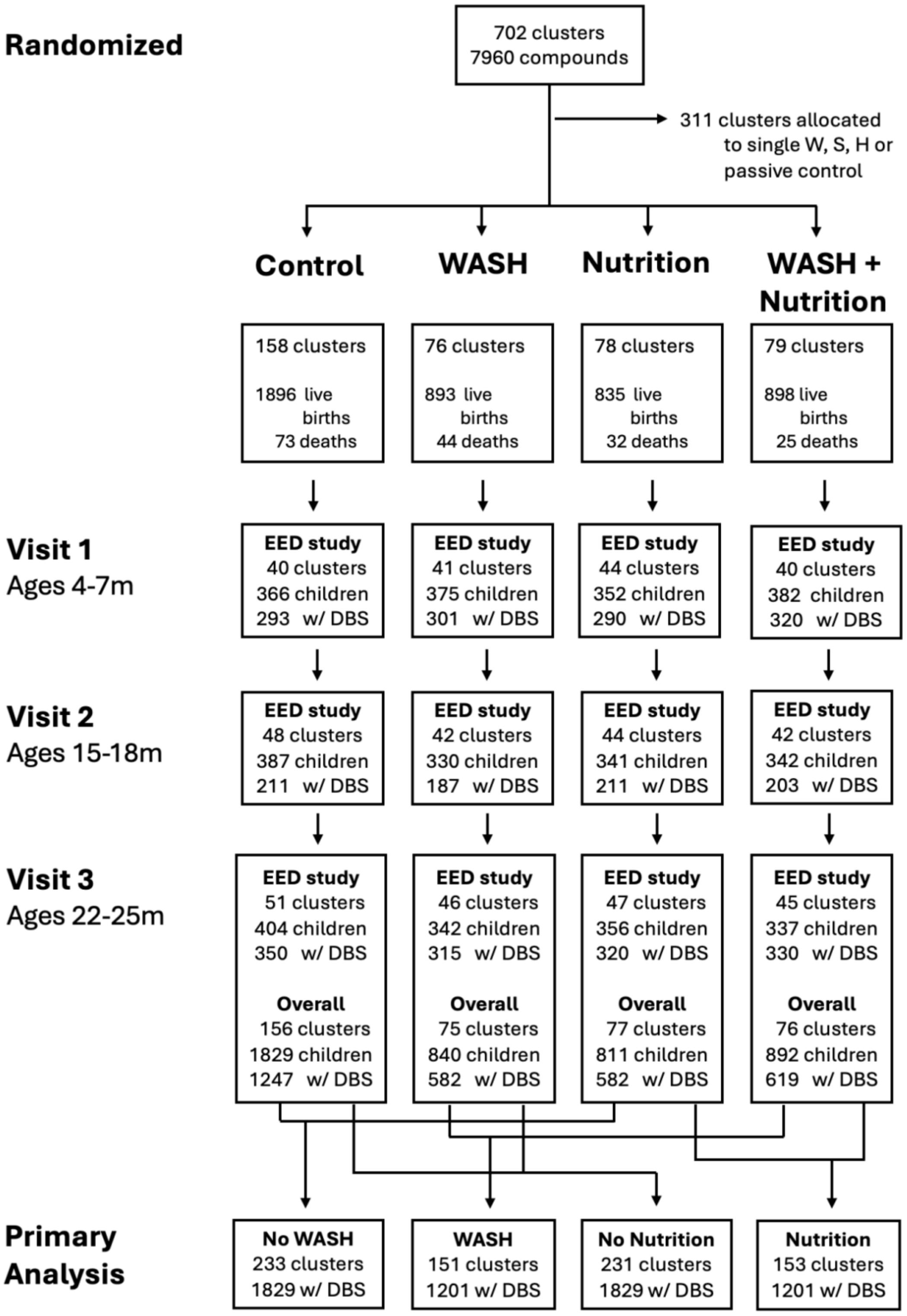

**Supplementary Table 3. Baseline characteristics by intervention group in the WASH Benefits Kenya trial.** Data are mean (SD) or n (%). Percentages are calculated among participants with nonmissing data for each characteristic.

| Characteristic | Control (N=1247) | WASH (N=582) | Nutrition (N=582) | Nutrition + WASH (N=619) |
|---|---|---|---|---|
| **Maternal** | | | | |
| Age (years) | 26.5 (6.3) | 26.6 (6.2) | 26.5 (6.2) | 26.5 (6.4) |
| Completed at least primary education | 589 (47%) | 261 (45%) | 275 (47%) | 289 (47%) |
| Height (cm) | 160 (5.9) | 160 (5.8) | 160 (5.8) | 160 (6) |
| Study child is firstborn | 232 (19%) | 94 (16%) | 124 (21%) | 122 (20%) |
| **Paternal** | | | | |
| Completed at least primary education | 707 (61%) | 338 (61%) | 350 (64%) | 348 (60%) |
| Works in agriculture | 526 (44%) | 257 (46%) | 250 (45%) | 270 (45%) |
| **Household** | | | | |
| Number of households per compound | 1.6 (1.1) | 1.7 (1.1) | 1.7 (1.2) | 1.7 (1.2) |
| Number of people per compound | 8.2 (5.4) | 8.3 (5.2) | 8.3 (6.1) | 8.6 (5.5) |
| Number of children <18 years in household | 2.9 (2) | 2.8 (2) | 2.9 (2) | 3.1 (2.1) |
| Has electricity | 77 (6%) | 39 (7%) | 46 (8%) | 50 (8%) |
| Has a cement floor | 62 (5%) | 29 (5%) | 29 (5%) | 32 (5%) |
| Owns a mobile phone | 1001 (80%) | 468 (80%) | 479 (82%) | 501 (81%) |
| Owns a motorcycle | 128 (10%) | 37 (6%) | 52 (9%) | 52 (8%) |
| **Drinking water** | | | | |
| Primary drinking water source is improved* | 934 (75%) | 408 (70%) | 425 (73%) | 472 (77%) |
| One-way walking time to primary water source (min) | 10.5 (11.5) | 11 (13.3) | 11.3 (11.6) | 10.7 (12.3) |
| Reported treating stored water | 128 (12%) | 65 (14%) | 56 (12%) | 70 (14%) |
| **Sanitation** | | | | |
| Own any latrine | 1035 (83%) | 494 (85%) | 493 (85%) | 522 (85%) |
| Access to improved latrine | 200 (17%) | 92 (17%) | 81 (15%) | 83 (14%) |
| Human faeces observed in the compound | 104 (8%) | 48 (8%) | 57 (10%) | 61 (10%) |

| | | | | |
|---|---|---|---|---|
| **Handwashing location** | | | | |
| Has water within 2 m of handwashing location | 312 (25%) | 156 (27%) | 145 (25%) | 167 (27%) |
| Has soap within 2 m of handwashing location | 97 (8%) | 80 (14%) | 57 (10%) | 69 (11%) |
| **Food security** | | | | |
| Prevalence of moderate-to-severe household hunger | 124 (10%) | 63 (11%) | 63 (11%) | 69 (11%) |

**Supplementary Table 4. Conditional and marginal seroprevalence by pathogen at 24 month-visit in in the WASH Benefits Kenya trial. Median (IQR) age at the time of measurement was 23 (22, 25) months.**

| Prevalence by treatment group | | Marginal prevalence, combining groups | | Risk difference (95% CI) | P value |
|---|---|---|---|---|---|
| **Group** | **n/N (%)** | **Group** | **n/N (%)** | | |
| **Viruses** | | | | | |
| **Norovirus GI/GII** | | | | | |
| Control | 1171/1247 (93.9%) | No WASH | 1715/1829 (93.8%) | Ref | - |
| WASH | 550/582 (94.5%) | WASH | 1131/1201 (94.2%) | 0.4 (-1.2, 2.1) | 0.63 |
| Nutrition | 544/582 (93.5%) | No Nutrition | 1721/1829 (94.1%) | Ref | - |
| WASH + Nutrition | 581/619 (93.9%) | Nutrition | 1125/1201 (93.7%) | -0.8 (-2.4, 0.8) | 0.39 |
| **Bacteria** | | | | | |
| ***Campylobacter* (p18, p39)** | | | | | |
| Control | 1204/1247 (96.6%) | No WASH | 1766/1829 (96.6%) | Ref | - |
| WASH | 570/582 (97.9%) | WASH | 1164/1201 (96.9%) | 0.7 (-0.5, 1.9) | 0.33 |
| Nutrition | 562/582 (96.6%) | No Nutrition | 1774/1829 (97%) | Ref | - |
| WASH + Nutrition | 594/619 (96%) | Nutrition | 1156/1201 (96.3%) | -1.1 (-2.3, 0.2) | 0.14 |
| ***V. cholerae* (OSP-Ogawa, OSP-Inaba)** | | | | | |
| Control | 22/1246 (1.8%) | No WASH | 27/1828 (1.5%) | Ref | - |
| WASH | 11/582 (1.9%) | WASH | 17/1201 (1.4%) | 0.3 (-0.5, 1.0) | 0.58 |
| Nutrition | 5/582 (0.9%) | No Nutrition | 33/1828 (1.8%) | Ref | - |
| WASH + Nutrition | 6/619 (1%) | Nutrition | 11/1201 (0.9%) | -1.1 (-1.8, -0.3) | 0.02 |

| | | | | | |
|---|---|---|---|---|---|
| **LT-ETEC (LTB)** | | | | | |
| Control | 1243/1246 (99.8%) | No WASH | 1823/1828 (99.7%) | Ref | - |
| WASH | 582/582 (100%) | WASH | 1200/1201 (99.9%) | 0.2 (0.0, 0.5) | 0.21 |
| Nutrition | 580/582 (99.7%) | No Nutrition | 1825/1828 (99.8%) | Ref | - |
| WASH + Nutrition | 618/619 (99.8%) | Nutrition | 1198/1201 (99.8%) | -0.1 (-0.4, 0.2) | 0.75 |
| **S. enterica typhi (HlyE)** | | | | | |
| Control | 280/1246 (22.5%) | No WASH | 424/1828 (23.2%) | Ref | - |
| WASH | 132/582 (22.7%) | WASH | 291/1201 (24.2%) | 0.3 (-2.6, 3.2) | 0.88 |
| Nutrition | 144/582 (24.7%) | No Nutrition | 412/1828 (22.5%) | Ref | - |
| WASH + Nutrition | 159/619 (25.7%) | Nutrition | 303/1201 (25.2%) | 2.6 (-0.3, 5.5) | 0.14 |
| **S. enterica (LPS B,D)** | | | | | |
| Control | 88/1246 (7.1%) | No WASH | 129/1828 (7.1%) | Ref | - |
| WASH | 36/582 (6.2%) | WASH | 88/1201 (7.3%) | 0.1 (-1.6, 1.9) | 0.92 |
| Nutrition | 41/582 (7%) | No Nutrition | 124/1828 (6.8%) | Ref | - |
| WASH + Nutrition | 52/619 (8.4%) | Nutrition | 93/1201 (7.7%) | 0.5 (-1.3, 2.3) | 0.63 |
| **Shigella/EIEC (IpaB)** | | | | | |
| Control | 1039/1246 (83.4%) | No WASH | 1516/1828 (82.9%) | Ref | - |
| WASH | 479/582 (82.3%) | WASH | 970/1201 (80.8%) | -2.0 (-4.6, 0.7) | 0.21 |
| Nutrition | 477/582 (82%) | No Nutrition | 1518/1828 (83%) | Ref | - |
| WASH + Nutrition | 491/619 (79.3%) | Nutrition | 968/1201 (80.6%) | -2.7 (-5.5, 0.0) | 0.08 |

| | | | | | |
|---|---|---|---|---|---|
| **Protozoans** | | | | | |
| **Cryptosporidium (Cp17, Cp23)** | | | | | |
| Control | 989/1247 (79.3%) | No WASH | 1456/1829 (79.6%) | Ref | - |
| WASH | 462/582 (79.4%) | WASH | 955/1201 (79.5%) | -0.4 (-3.2, 2.5) | 0.82 |
| Nutrition | 467/582 (80.2%) | No Nutrition | 1451/1829 (79.3%) | Ref | - |
| WASH + Nutrition | 493/619 (79.6%) | Nutrition | 960/1201 (79.9%) | 0.5 (-2.5, 3.4) | 0.77 |
| **Giardia (VSP-5)** | | | | | |
| Control | 629/1246 (50.5%) | No WASH | 908/1828 (49.7%) | Ref | - |
| WASH | 304/582 (52.2%) | WASH | 626/1201 (52.1%) | 2.6 (-1.3, 6.5) | 0.25 |
| Nutrition | 279/582 (47.9%) | No Nutrition | 933/1828 (51%) | Ref | - |
| WASH + Nutrition | 322/619 (52%) | Nutrition | 601/1201 (50%) | -1.1 (-5.0, 2.7) | 0.61 |
| **E. histolytica (LecA)** | | | | | |
| Control | 113/1246 (9.1%) | No WASH | 183/1828 (10%) | Ref | - |
| WASH | 75/582 (12.9%) | WASH | 131/1201 (10.9%) | 0.4 (-1.9, 2.8) | 0.75 |
| Nutrition | 70/582 (12%) | No Nutrition | 188/1828 (10.3%) | Ref | - |
| WASH + Nutrition | 56/619 (9%) | Nutrition | 126/1201 (10.5%) | 0.4 (-1.9, 2.8) | 0.75 |
| **Helminths** | | | | | |
| **T. solium (Es33, T24)** | | | | | |
| Control | 149/1246 (12%) | No WASH | 215/1828 (11.8%) | Ref | - |
| WASH | 53/582 (9.1%) | WASH | 129/1201 (10.7%) | -1.4 (-3.7, 0.9) | 0.30 |
| Nutrition | 66/582 (11.3%) | No Nutrition | 202/1828 (11.1%) | Ref | - |

| | | | | | |
|---|---|---|---|---|---|
| WASH + Nutrition | 76/619 (12.3%) | Nutrition | 142/1201 (11.8%) | 0.6 (-1.7, 2.9) | 0.66 |
| **S. stercoralis (NIE)** | | | | | |
| Control | 13/1246 (1%) | No WASH | 18/1828 (1%) | Ref | - |
| WASH | 6/582 (1%) | WASH | 12/1201 (1%) | 0.0 (-0.6, 0.7) | 0.96 |
| Nutrition | 5/582 (0.9%) | No Nutrition | 19/1828 (1%) | Ref | - |
| WASH + Nutrition | 6/619 (1%) | Nutrition | 11/1201 (0.9%) | -0.1 (-0.8, 0.5) | 0.69 |

**Supplementary Figure 5. Age-dependent seroprevalence for enteric pathogens by nutrition treatment in the WASH Benefits Kenya trial. A.** Higher transmission pathogens. **B.** Lower transmission pathogens. The only difference between panels is the Y-axis range, to better show the range of the data. Lines age-seroprevalence estimated by treatment group using locally weighted regression over individual data (points), trimmed to ages 4-28 months to avoid edge effects. Points and error bars to the right of each panel show treatment-specific marginal average seroprevalence and 95% confidence intervals by study visit (1, 2, 3). Gray shading shows child age ranges of visits 1 and 3.

**A**

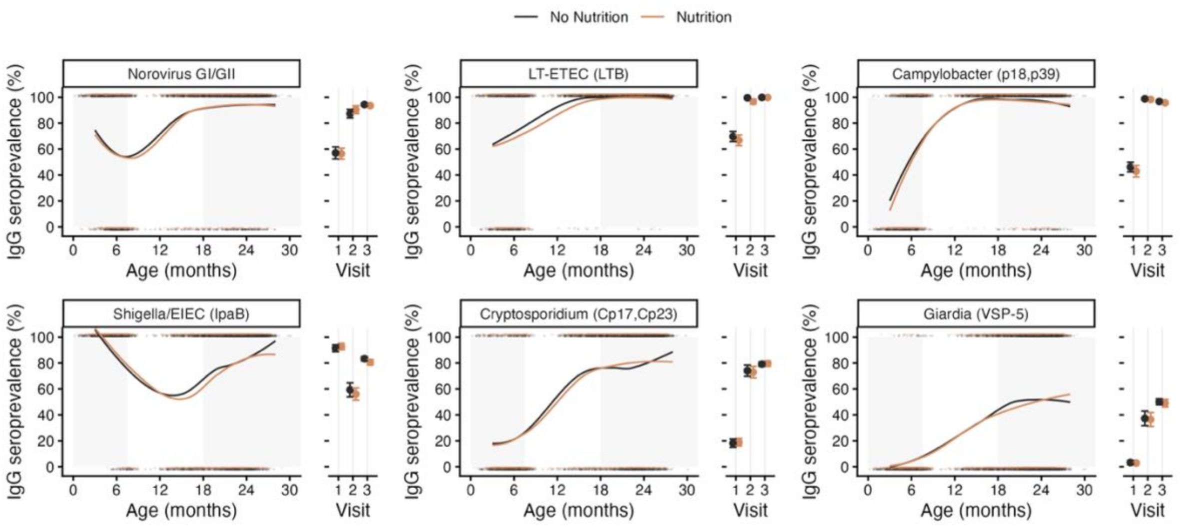


**B**

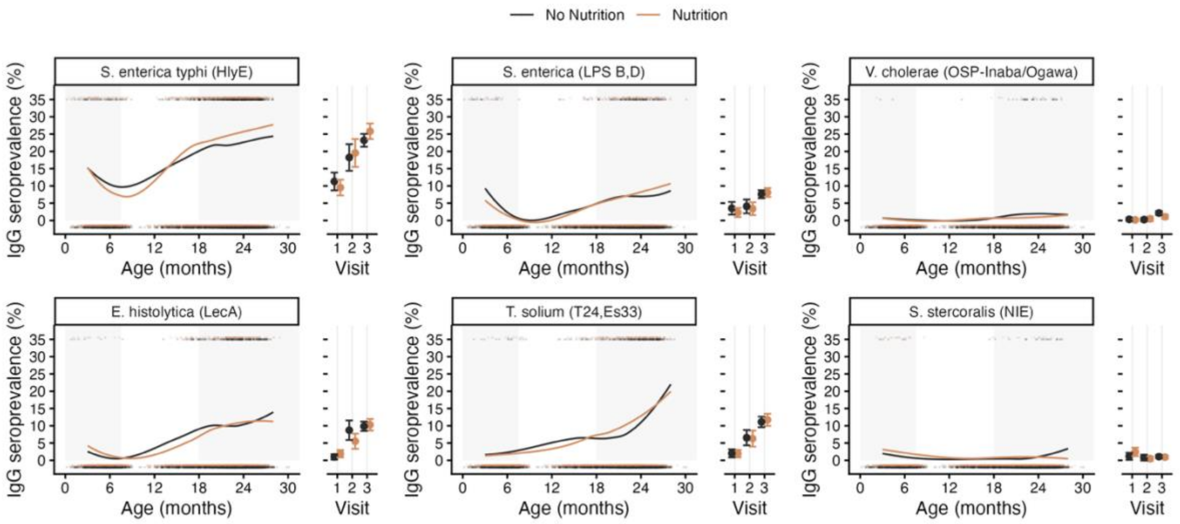

**Supplementary Figure 6. Distribution of the number of enteric pathogens to which children were seropositive at the 24-month visit, by WASH and nutrition intervention assignment in the WASH Benefits Kenya trial.**

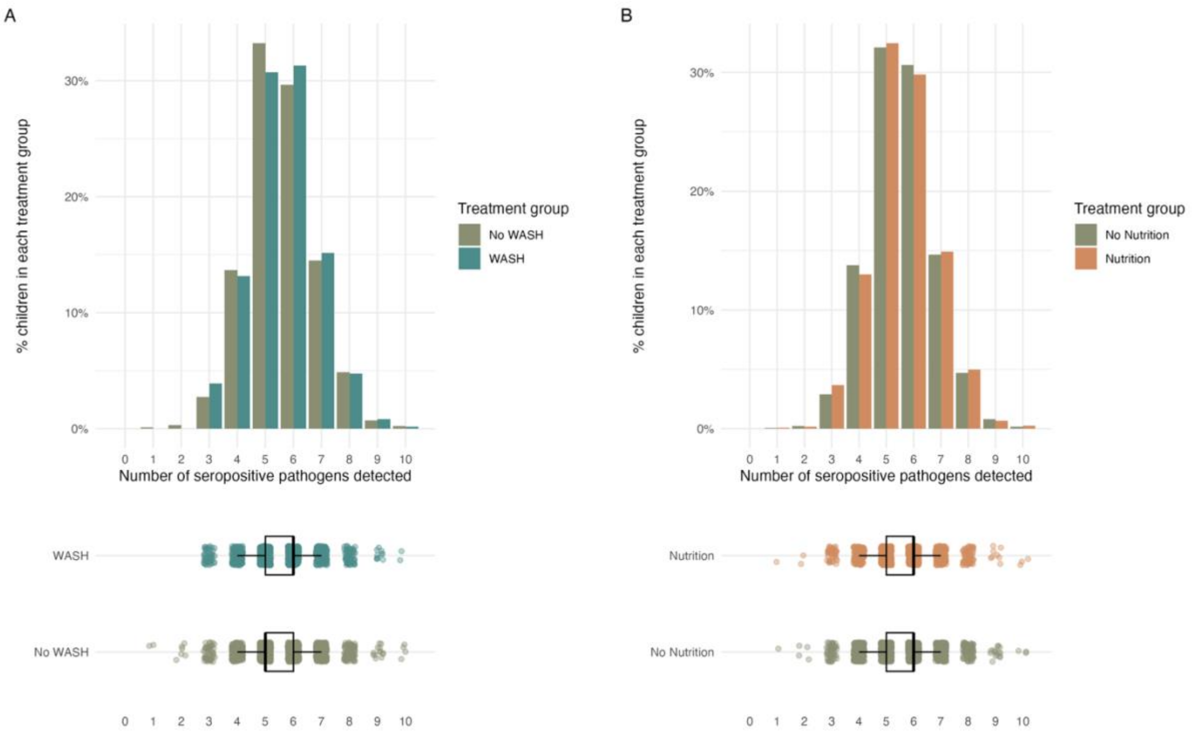

**Supplementary Table 7.** Intervention adherence at enrollment and at 1- and 2-year follow-up by 2 x 2 factorial WASH and Nutrition margins in the WASH Benefits Kenya trial.

| Indicator | Round | WASH factorial margin | | Nutrition factorial margin | |
|---|---|---|---|---|---|
| | | WASH | No WASH | Nutrition | No Nutrition |
| Number of compounds assessed | Enrollment | 1830/1833 (100%) | 2756/2762 (100%) | 1761/1764 (100%) | 2825/2831 (100%) |
| | Year 1 | 1029/1833 (56%) | 1497/2762 (54%) | 947/1764 (54%) | 1579/2831 (56%) |
| | Year 2 | 1410/1833 (77%) | 2108/2762 (76%) | 1385/1764 (79%) | 2133/2831 (75%) |
| Visited by promoter in past month | Enrollment | .. | .. | .. | .. |
| | Year 1 | 774/986 (78%) | 1010/1413 (71%) | 732/907 (81%) | 1052/1492 (71%) |
| | Year 2 | 500/1359 (37%) | 743/2047 (36%) | 510/1345 (38%) | 733/2061 (36%) |
| Stored drinking water has detectable free chlorine | Enrollment | 55/1440 (4%) | 58/2190 (3%) | 40/1390 (3%) | 73/2240 (3%) |
| | Year 1 | 336/791 (42%) | 34/1239 (3%) | 165/759 (22%) | 205/1271 (16%) |
| | Year 2 | 240/1250 (19%) | 53/1979 (3%) | 143/1266 (11%) | 150/1963 (8%) |
| Access to improved latrine | Enrollment | 296/1739 (17%) | 428/2582 (17%) | 262/1666 (16%) | 462/2655 (17%) |
| | Year 1 | 897/1003 (89%) | 241/1417 (17%) | 488/901 (54%) | 650/1519 (43%) |
| | Year 2 | 1090/1350 (81%) | 370/1994 (19%) | 660/1319 (50%) | 800/2025 (40%) |
| Child faeces safely disposed of | Enrollment | 121/722 (17%) | 147/1031 (14%) | 89/663 (13%) | 179/1090 (16%) |
| | Year 1 | 613/895 (68%) | 493/1294 (38%) | 442/823 (54%) | 664/1366 (49%) |
| | Year 2 | 424/1254 (34%) | 188/1898 (10%) | 271/1235 (22%) | 341/1917 (18%) |
| Handwashing location has water and soap | Enrollment | 117/1830 (6%) | 153/2756 (6%) | 110/1761 (6%) | 160/2825 (6%) |
| | Year 1 | 797/1029 (77%) | 185/1497 (12%) | 442/947 (47%) | 540/1579 (34%) |
| | Year 2 | 282/1410 (20%) | 203/2108 (10%) | 228/1385 (16%) | 257/2133 (12%) |
| LNS sachets consumed (% expected)* | Enrollment | - | - | - | - |
| | Year 1 | 96% | 95% | 95% | - |
| | Year 2 | 116% | 114% | 115% | - |

*Note.* Data are n/N (%) or %. "-" indicates that a measure was not collected or was not interpretable for that factorial margin. Free chlorine was measured in every trial arm. *LNS consumption was reported for the Nutrition group only and is conditional on a child being in a Nutrition cluster, expressed as a percentage of expected consumption. Promoter visits and LNS consumption were not measured at enrollment. WASH = water, sanitation, and handwashing; LNS = lipid-based nutrient supplement.
